# Modelling 25-hydroxyvitamin D responses to different recommended daily intakes of vitamin D

**DOI:** 10.1101/2023.06.16.23291505

**Authors:** Tao You, Nadda Muhamad, Joseph Jenner, Zhonghui Huang

## Abstract

The reference nutrient intake for vitamin D in people aged ≥ 4 years is 10 μg/day in the UK, which contrasts with the recommended daily allowance of 15 μg/day for people aged 1-70 years in the USA. Using modelling of published data, we compared 95% confidence intervals for serum concentrations of 25-hydroxyvitamin D_3_ (25(OH)D_3_) attained in response to these two doses. 97.5% of those taking 10 μg/day vs. 15 μg/day were predicted to attain circulating 25(OH)D concentrations >44 nmol/L vs. 50 nmol/L, respectively.

## Main text

Vitamin D is crucial for maintaining bone mineralisation, which helps prevent osteopenia and osteoporosis (Holick, 2007).

Vitamin D is synthesised in the skin by exposure to ultraviolet B (UVB) radiation. Its major circulating metabolite, 25-hydroxyvitamind D (i.e. 25(OH)D), is subsequently activated to form 1,25(OH)2D (calcitriol) which exerts its biological effects. Vitamin D deficiency is prevalent in the UK and USA, especially in spring and winter (Hyppönen & Power, 2007; Ganji *et al*, 2012; Lin *et al*, 2020).

Vitamin D status is characterised by the serum levels of 25(OH)D. Thresholds are controversial and vary internationally (Dominguez *et al*, 2021). According to the UK Department of Health, serum 25(OH)D should not be lower than 25 nmol/L (SACN, 2016). However, deleterious effects were reported to be associated with serum 25(OH)D < 50 nmol/L (Malabanan *et al*, 1998; Gaksch et al, 2017; Giustina *et al*, 2020; Griffin *et al*, 2021). Others have proposed 75 nmol/L or even higher concentrations as a sufficiency target (Holick, 2007, Dominguez *et al*, 2021).

The UK Scientific Advisory Committee on Nutrition (SACN) recommends a reference nutrient intake (RNI) of 10 µg/day vitamin D in winter and spring for the general population and 10 µg/day all year round for the at-risk groups (SACN, 2016). On the other hand, the US National Academy of Medicine (NAM) recommends a daily allowance of 15 µg/day vitamin D for 1-to-70-year-old, and 20 µg/day for over 70s (Bouillon, 2017). Here, we investigate whether there is significant difference between 10 and 15 µg/day doses in terms of serum 25(OH)D PK profiles, and how they compare with the 25, 50 and 75 nmol/L thresholds for 25(OH)D proposed in the field.

To answer this question, we first searched the literature for clinical trials reporting effects of a daily dose of 10 µg vitamin D_3_ dose either with or without calcium or other vitamins in participants with baseline 25(OH)D concentrations <25 nmol/L. We limited the dosage form to tablets, chewable tablets, pills and capsules and found 4 arms of 3 randomised controlled trials (RCTs) between 1979 and 2019 (Lips *et al*., 1988; Chel *et al*., 1998; Grados *et al*., 2003). All studies focused on elderly subjects and bodyweight was not reported. The mean serum 25(OH)D reached 60 nmol/L after 12 weeks (dots with solid lines in Figure 2A). Similarly, 2 arms of 2 RCTs for 15 µg/day vitamin D_3_ dose were identified (dots with solid lines in Figure 2B): both studies focused on elderly subjects, and bodyweight was unavailable (Chel *et al*., 1998; Heijboer *et al*., 2015).

Next, we used modelling and simulation to generate the 95% confidence interval (CI) for the serum 25(OH)D concentrations attained using this dose, irrespective of bodyweight.

Previously, we reported a physiologically-based pharmacokinetic (PBPK) model for serum pharmacokinetics (PK) of vitamin D_3_ and 25(OH)D_3_ (Figure 1). This naïve averaged model employed only a single set of parameters and made accurate predictions for adult populations from Asia, Europe, Americas, and Oceania in all seasons under a wide range of doses (repeated daily dosing: 83 clinical trial arms, 12.5 – 1250 µg/day; large single doses: 16 clinical trial arms, 1250 – 50000 µg) (Huang & You, 2021).

**Figure 1.**
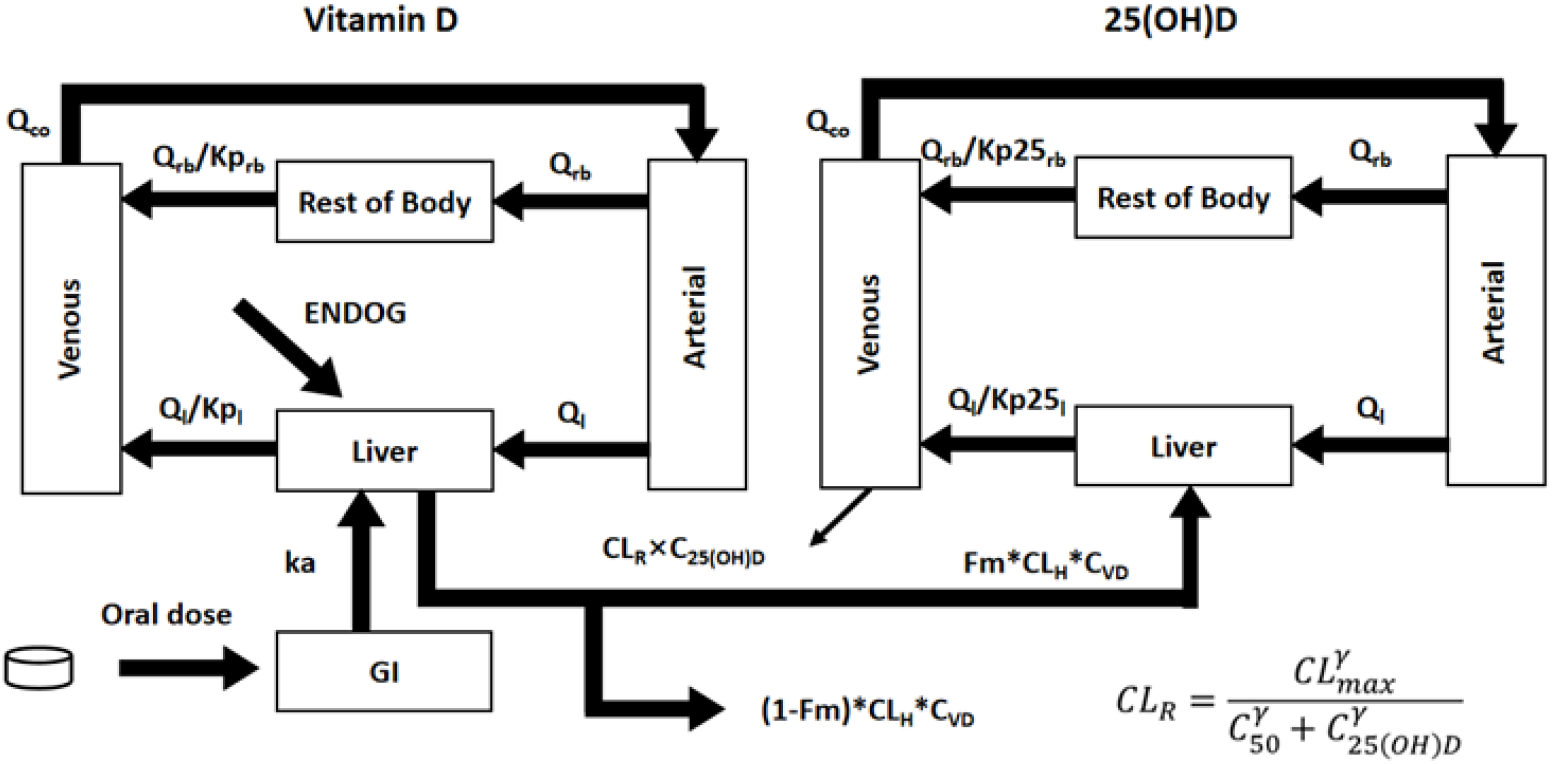
Model diagram. Reproduced from Huang & You, 2021. ENDOG, endogenous vitamin D production rate, GI, gastrointestinal compartment.

Here, we further developed it into a nonlinear mixed effects (NLME) model. We first refitted the vitamin D part of the model with the same data from (Huang & You, 2021) (13 arms: single dose between 70 – 2500 µg; repeated dose between 20 – 275 µg/day). The best model inherited all parameters from the naïve average model but the hepatic clearance parameter CL_H_: for repeated daily dosing, NLME: 0.222 h ^-1^, 95% CI [0.179, 0.274]; naïve average: h ^-1^ (Table 1: “Vitamin D Part”. Also referred to as model 11 Table S5).

**Table 1.**
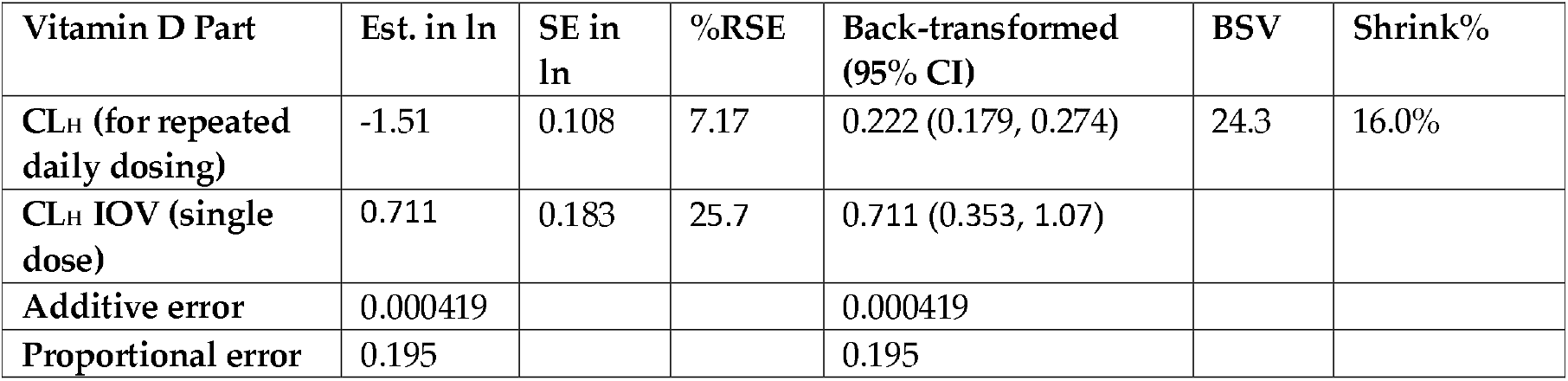
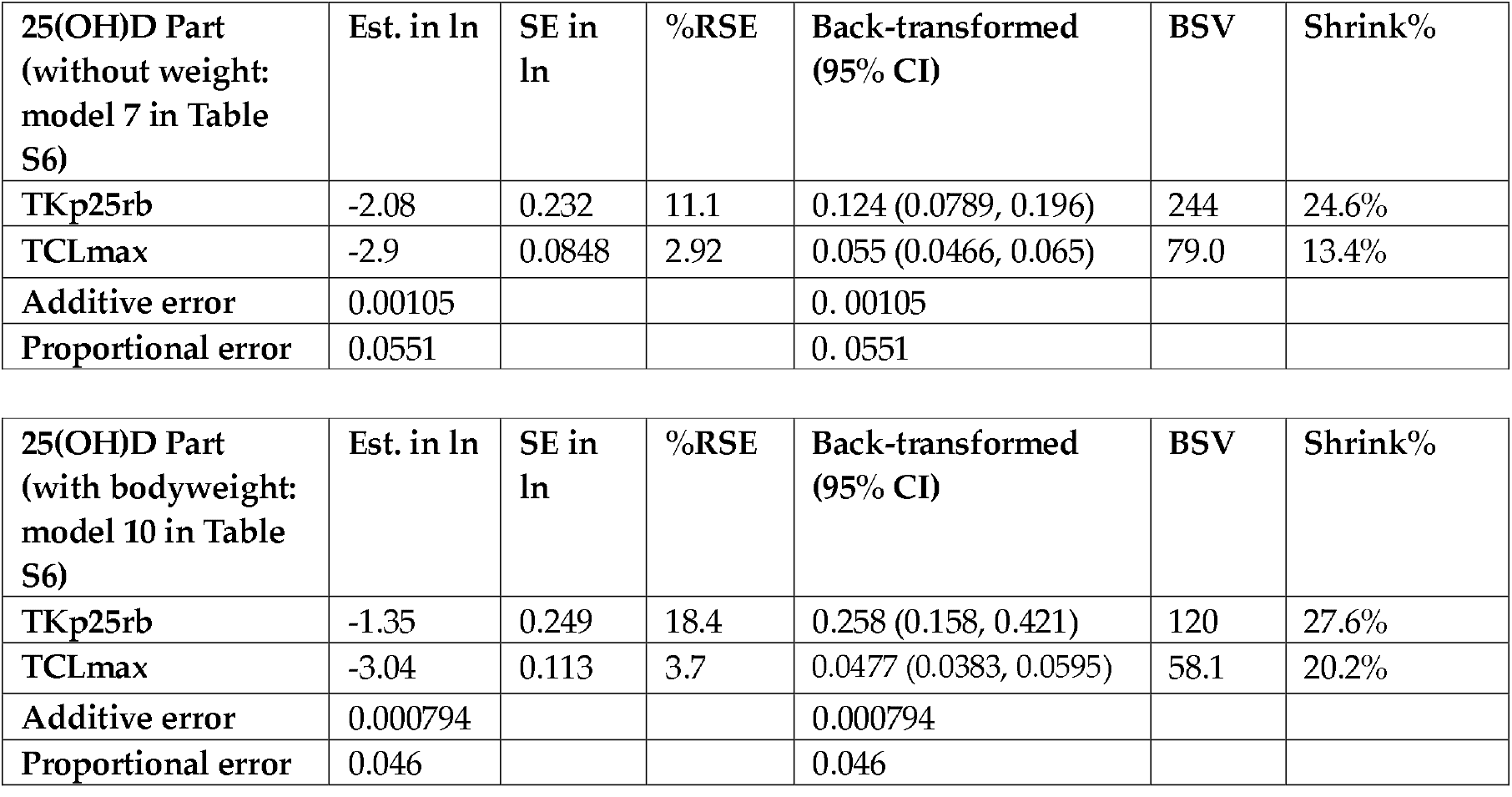
Parameters for the final model. Models were fitted using the SAEM method in nlmixr R package.

| Vitamin D Part | Est. in ln | SE in ln | %RSE | Back-transformed (95% CI) | BSV | Shrink% |
| --- | --- | --- | --- | --- | --- | --- |
| $CL_H$ (for repeated daily dosing) | -1.51 | 0.108 | 7.17 | 0.222 (0.179, 0.274) | 24.3 | 16.0% |
| $CL_H$ IOV (single dose) | 0.711 | 0.183 | 25.7 | 0.711 (0.353, 1.07) | | |
| Additive error | 0.000419 |  |  | 0.000419 |  |  |
| Proportional error | 0.195 |  |  | 0.195 |  |  |

| 25(OH)D Part<br>(without weight:<br>model 7 in Table<br>S6) | Est. in ln | SE in<br>ln | %RSE | Back-transformed<br>(95% CI) | BSV | Shrink% |
| --- | --- | --- | --- | --- | --- | --- |
| TKp25rb | -2.08 | 0.232 | 11.1 | 0.124 (0.0789, 0.196) | 244 | 24.6% |
| TCLmax | -2.9 | 0.0848 | 2.92 | 0.055 (0.0466, 0.065) | 79.0 | 13.4% |
| Additive error | 0.00105 |  |  | 0.00105 |  |  |
| Proportional error | 0.0551 |  |  | 0.0551 |  |  |

| 25(OH)D Part<br>(with bodyweight:<br>model 10 in Table<br>S6) | Est. in ln | SE in<br>ln | %RSE | Back-transformed<br>(95% CI) | BSV | Shrink% |
| --- | --- | --- | --- | --- | --- | --- |
| TKp25rb | -1.35 | 0.249 | 18.4 | 0.258 (0.158, 0.421) | 120 | 27.6% |
| TCLmax | -3.04 | 0.113 | 3.7 | 0.0477 (0.0383, 0.0595) | 58.1 | 20.2% |
| Additive error | 0.000794 |  |  | 0.000794 |  |  |
| Proportional error | 0.046 |  |  | 0.046 |  |  |

We then constructed a 90-day 25(OH)D training set for the sake of computing time (90 arms in total; doses from 10 to 1250 µg/day) and refitted the 25(OH)D part of the model based on the expected parameter values for the vitamin D part. Our final model for the 25(OH)D part includes random effects for the partition coefficient Kp_25rb_ and maximum clearance CL_max_, does not have any covariates, and makes good parametric inference (Table 1: “25(OH)D Part (without weight)”. Also referred to as model 7 in Table S6). It is important to note the training data span a wide variety of race, age, gender, and geographical location, as mentioned earlier. Hence, the model is not limited by these factors and can be viewed as a general model for any population in the world.

We then generated the 95% CI for attained serum 25(OH)D for the 10 µg/day dose by sampling the NLME model. Encouragingly, all reported mean values were within the 95% CI (Figure 2A). We found the baseline 25(OH)D levels between 18 and 24 nmol/L only had limited effects on the predicted serum 25(OH)D concentrations at 26 weeks (dashed lines in Figure 2A) ranging from 44 to 72 nmol/L. This is the consistent with the observations that people with the lowest baseline levels had the highest increment in 25(OH)D in response to the same dose (Huang & You, 2021). Given the lower bound of 44 nmol/L, model predicts >2.5% chance for a person with baseline 25(OH)D < 25 nmol/L to fall below the 50 nmol/L threshold at the 10 µg/day dose.

**Figure 2.**
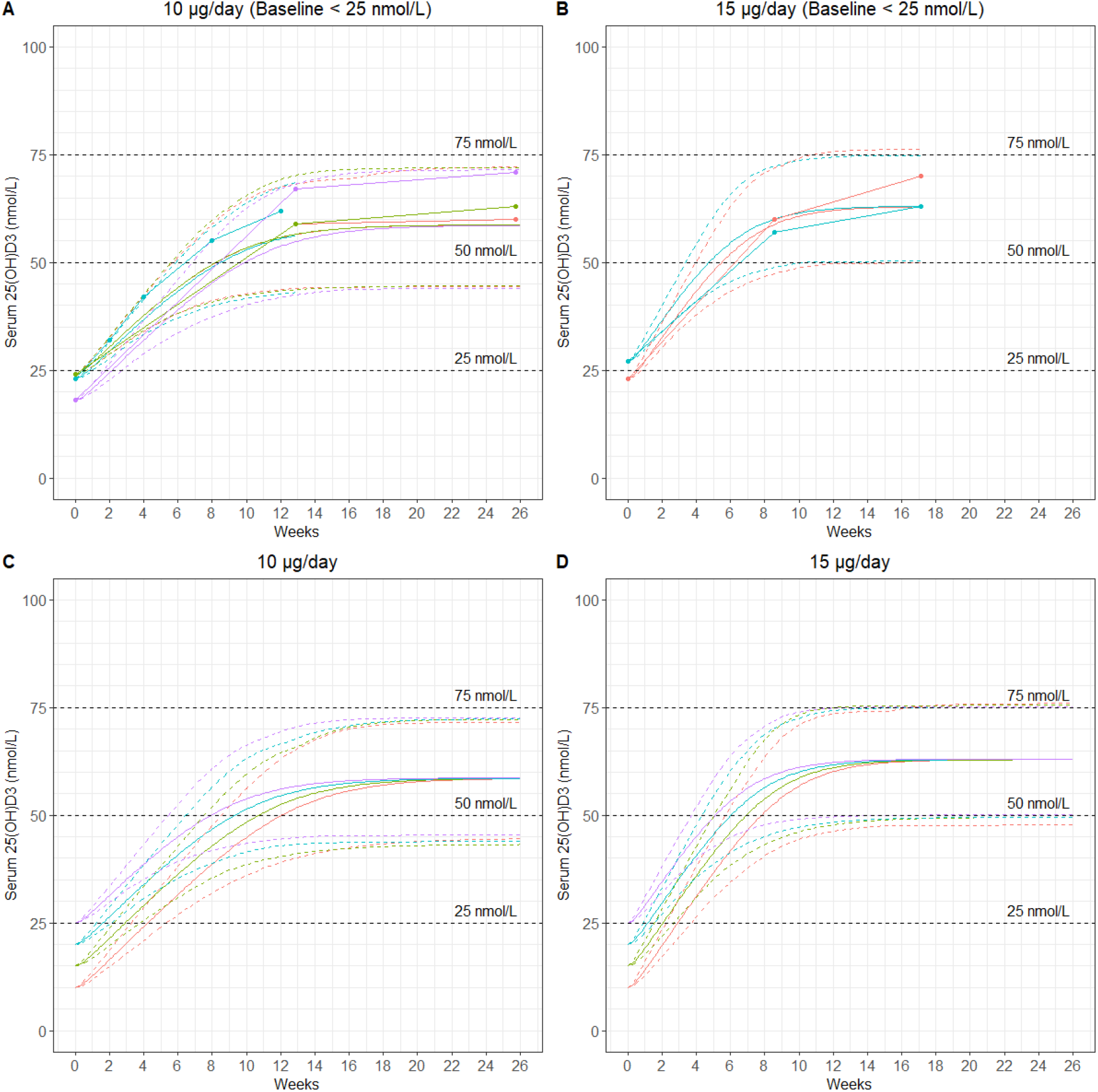
Simulation of the repeated dosing at 10 µg/day (Figure 1A: baseline 25(OH)D at 18, 23 and 24 nmol/L) and 15 µg/day (Figure 1B: baseline 25(OH)D at 23 or 27 nmol/L). Simulation of serum 25(OH)D for repeated dosing at 10 (C) and 15 µg/day (D) for 26 weeks at each baseline level of 10 (red), 15 (green), 20 (blue) and 25 (purple) nmol/L. For sampling, 95% confidence interval was generated by sampling 1000 subjects using all parameters with the Latin Hypercube method. Mean (solid lines) and 95% confidence intervals (dashed lines) are shown.

We simulated the 15 µg/day dose for two trial arms with baseline values at 23 and 27 nmol/L, respectively (dots with solid lines in Figure 2B). All data points were within the predicted 95% CI (Figure 2B). Interestingly, the lower bound of the predicted 95% was 50 nmol/L (dashed curves in Figure 1B). This is consistent with the US NAM recommendation that 15 µg/day dose is needed to achieve the 50 nmol/L target, though the simulation is made for a person with baseline levels >23 nmol/L and the claim is made for the majority of the US population.

We next evaluated the impact of 25(OH)D baselines on predicted steady-state 25(OH)D levels by simulating different baseline values between 10 and 25 nmol/L. The predicted steady state serum 25(OH)D concentration was insensitive to baseline values after approximately 18 weeks for 10 µg/day dose (Figure 2C) and 16 weeks for 15 µg/day dose (Figure 2D). In other words, the 25(OH)D baseline might be unimportant for the steady state, and these simulation results support the recommendation that 15 µg/day dose is sufficient for 97.5% of adult population to achieve the 50 nmol/L target.

Vitamin D is lipophilic and distributes into the fat tissue. We explored whether bodyweight or body mass index (BMI) could be incorporated into the model. Among the 25(OH)D data we used, 33 out of 90 arms reported mean bodyweight ([45, 93] kg). Our best model (model 10 in Table S6) does not consider BMI. Instead, it assumes volume of distribution for each compartment is proportional to bodyweight. For instance, *V*_*rb*_ = 62.6 × *WT*/70, where *V*_*rb*_ = 62.6*L* for a 70kg man. This new model reduces BSV for the 2 parameters Kp_25rb_ and CL_max_ (Table 1: “25(OH)D Part (with bodyweight)”) and moderately improves the goodness of fit. When we repeated the simulations in Figures 2C and 2D for subjects weighing 45kg and 93kg, the qualitative conclusions on the 2 doses remain the same (Figure S1).

In summary, based on a well-qualified naïve averaged model, we developed an NLME PBPK model to assess the 95% CI of 25(OH)D serum PK at 10 and 15 µg/day doses. Our simulation suggests repeated dosing at 15 µg/day is sufficient for 97.5% of population to reach the 50 nmol/L threshold, while 10 µg/day elevates 25(OH)D concentrations >44 nmol/L in 97.5% of people. This may be important to consider when choosing a daily dose of vitamin D to prevent vitamin D deficiency.

## Data Availability

All data produced in the present study are available upon reasonable request to the authors

## Contributions

Designed the research: TY. Collected data: ZHH. Collated data: NM, TY. Performed modelling: All authors. Analysed the data: All authors. Drafted the paper: TY. Finalised on the paper: All authors.

## Acknowledgement

We are grateful for the time and useful comments made by Prof Martin Hewison (University of Birmingham) and Prof Adrian R Martineau (Queen Mary University of London), which significantly improved the manuscript. The authors would also like to thank Prof Kesara Na-Bangchang for the constructive advice. NM, TY and Prof Kesara Na-Bangchang are supported by the Royal Golden Jubilee Ph.D. Programme in Thailand (grant number PHD/0095/2561).

