## Supplementary material for "Modelling 25-hydroxyvitamin D responses to different recommended daily intakes of vitamin D": Table S1

**Table S1.** Primary characteristics of vitamin D repeated daily dose studies on 25(OH)D PK.

| Ref No. | Arm No. | Year | Study Type | No. of participants | Drug | Dosage  form | Dose  (µg) | Dose  (IU) | Duration  (d) | Country | Age (year) | Women (%) | Weight (kg) | BMI (kg/m^2^) | SCR* (mg/dL) | Season^§^ (W/N) | Measurement |
| --- | --- | --- | --- | --- | --- | --- | --- | --- | --- | --- | --- | --- | --- | --- | --- | --- | --- |
| 1 | 1 | 1982 | Non-RCT | 9 | Vitamin D3 | arachis oil | 10 | 400 | 70 | UK | 21.2 | 22 | 50.2 | NA | NA | NA | HPLC |
| 2 | 2 | 1988 | RCT | 25 | Vitamin D3 | Tablets | 10 | 400 | 360 | Netherlands | 81 | 84 | NA | NA | 1.0 | N | CPBA |
| 2 | 3 | 1988 | RCT | 30 | Vitamin D3 | Tablets | 10 | 400 | 360 | Netherlands | 84 | 83 | NA | NA | 1.0 | N | CPBA |
| 3 | 9 | 1998 | RCT | 15 | Vitamin D3 | Tablets | 10 | 400 | 90 | Netherlands | 84.4 | 100 | NA | NA | 0.9 | NA | RIA |
| 4 | 11 | 2003 | RCT | 95 | Vitamin D3 | chewable tablets | 10 | 400 | 360 | France | 74 | 100 | NA | 26.8 | NA | N | Others |
| 5 | 12 | 2003 | RCT | 67 | Vitamin D3 | chewable tablets | 10 | 400 | 30 | Denmark | 75 | 60 | NA | NA | 1.0 | W | RIA |
| 6 | 13 | 2005 | Non-RCT | 163 | Vitamin D3 | NA | 10 | 400 | 90 | Japan | 67.3 | 98 | NA | 22 | NA | N | RIA |
| 7 | 15 | 2009 | RCT | 29 | Vitamin D3 | Pills | 10 | 400 | 90 | Brazil | 61.3 | 100 | 60.3 | 26.7 | NA | NA | RIA |
| 8 | 18 | 2011 | Non-RCT | 512 | Vitamin D3 | Tablets | 10 | 400 | 112 | Canada | NA | NA | NA | NA | NA | W | RIA |
| 8 | 19 | 2011 | Non-RCT | 169 | Vitamin D3 | Tablets | 10 | 400 | 112 | Canada | NA | NA | NA | NA | NA | W | RIA |
| 9 | 20 | 2012 | RCT | 27 | Vitamin D3 | Fish oil Capsules | 10 | 400 | 28 | Norway | 28 | 63.6 | NA | 23.7 | NA | W | RIA |
| 9 | 21 | 2012 | Non-RCT | 28 | Vitamin D3 | Multivitamin tablets | 10 | 400 | 28 | Norway | 28 | 63.6 | NA | 23.7 | NA | W | RIA |
| 10 | 23 | 2013 | RCT | 20 | Vitamin D3 | Tablets | 10 | 400 | 90 | Thailand | 36 | 85 | 56.9 | 22.4 | NA | N | LC-MS |
| 11 | 24 | 2013 | RCT | 84 | Vitamin D3 | Capsules | 10 | 400 | 360 | UK | 64.2 | 100 | 68.1 | 25.3 | NA | W | LC-MS |
| 12 | 25 | 2013 | RCT | 20 | Vitamin D3 | Liquid | 10 | 400 | 34 | Canada | 58.9 | 0 | NA | 27.9 | NA | N | LC-MS |
| 14 | 27 | 1977 | Non-RCT | 18 | Vitamin D3 | oil droplet | 12.5 | 500 | 14 | Canada | 83.4 | 100 | NA | NA | NA | NA | CPBA |
| 14 | 28 | 1977 | Non-RCT | 6 | Vitamin D3 | oil droplet | 12.5 | 500 | 14 | Canada | 35 | 50 | NA | NA | NA | NA | CPBA |
| 15 | 29 | 1977 | RCT | 8 | Vitamin D | Tablets | 12.5 | 500 | 120 | UK | NA | NA | NA | NA | NA | W | CPBA |
| 15 | 30 | 1977 | RCT | 8 | Vitamin D | Tablets | 12.5 | 500 | 120 | UK | NA | NA | NA | NA | NA | W | CPBA |
| 16 | 31 | 2008 | RCT | 18 | Vitamin D3 | Capsules | 12.5 | 500 | 77 | USA | 35.5 | 72.2 | NA | 31.7 | NA | W | LC-MS |
| 17 | 32 | 2013 | RCT | 24 | Vitamin D3 | Tablets | 12.5 | 500 | 90 | India | NA | NA | NA | NA | NA | N | NA |
| 18 | 33 | 2007 | RCT | 55 | Vitamin D3 | Tablets | 15 | 600 | 120 | Netherlands | 84.3 | 83.6 | NA | NA | 1.0 | N | RIA |
| 19 | 35 | 2015 | RCT | 22 | Vitamin D3 | Tablets | 15 | 600 | 120 | Netherlands | 82 | 0 | NA | NA | NA | N | RIA |
| 20 | 42 | 1996 | Non-RCT | 41 | Vitamin D3 | Capsules | 20 | 800 | 63 | Netherlands | 75.3 | 100 | NA | NA | NA | W | CPBA |
| 20 | 43 | 1996 | Non-RCT | 6 | Vitamin D3 | Capsules | 20 | 800 | 28 | Netherlands | 30.1 | 100 | NA | NA | NA | W | CPBA |
| 21 | 44 | 1999 | Non-RCT | 10 | Vitamin D3 | Tablets | 20 | 800 | 84 | USA | 70 | 100 | 69.7 | 27.4 | NA | NA | Others |
| 22 | 45 | 2000 | RCT | 79 | Vitamin D3 | Tablets | 20 | 800 | 720 | UK | 59.2 | 100 | 62.4 | 24.1 | NA | N | RIA |
| 23 | 46 | 2001 | RCT | 35 | Vitamin D3 | NA | 20 | 800 | 360 | UK | 45.3 | 100 | 67.6 | 25.1 | NA | N | RIA |
| 23 | 47 | 2001 | RCT | 35 | Vitamin D3 | NA | 20 | 800 | 360 | UK | 49.1 | 100 | 67.4 | 25 | NA | N | RIA |
| 24 | 48 | 2002 | RCT | 13 | Vitamin D3 | Tablets | 20 | 800 | 56 | USA | 28.7 | 0 | NA | 25 | NA | W | CPBA |
| 24 | 49 | 2002 | RCT | 14 | Vitamin D3 | Tablets | 20 | 800 | 56 | USA | 72.8 | 0 | NA | 29 | NA | W | CPBA |
| 25 | 50 | 2003 | RCT | 62 | Vitamin D3 | Tablets | 20 | 800 | 90 | Switzerland | 84.9 | 100 | 60.5 | 24.7 | NA | W | RIA |
| 26 | 51 | 2007 | RCT | 104 | Vitamin D3 | Capsules | 20 | 800 | 90 | USA | 59.9 | 100 | 78 | 29 | NA | N | RIA |
| 27 | 52 | 2009 | RCT | 32 | Vitamin D3 | Gelatinous form | 20 | 800 | 30 | Japan | 83.8 | 75 | 45 | 20.2 | NA | N | RIA |
| 28 | 53 | 2010 | RCT | 72 | Vitamin D | Capsules | 20 | 800 | 180 | Netherlands | 40.5 | 75 | NA | 28.9 | NA | N | RIA |
| 29 | 54 | 2011 | RCT | 10 | Vitamin D3 | Capsules | 20 | 800 | 120 | Switzerland | 63.45 | 100 | NA | 25.49 | NA | NA | LC-MS |
| 30 | 56 | 2012 | RCT | 13 | Vitamin D3 | Capsules | 20 | 800 | 70 | Ireland | 57.2 | 61.5 | 79 | 28.3 | NA | W | ELISA |
| 13 | 57 | 2015 | RCT | 15 | Vitamin D3 | Capsules | 20 | 800 | 112 | China | 31.8 | 53 | NA | 22.8 | NA | W | LC-MS |
| 31 | 58 | 2015 | RCT | 14 | Vitamin D3 | Tablets | 20 | 800 | 182 | Netherlands | 85.5 | 57 | 69 | Normal | NA | N | CHEMI |
| 1 | 60 | 1982 | Non-RCT | 9 | Vitamin D3 | arachis oil | 25 | 1000 | 70 | UK | 21.4 | 22.3 | 45.9 | Low | NA | NA | HPLC |
| 32 | 63 | 1998 | RCT | 13 | Vitamin D3 | Capsules | 25 | 1000 | 56 | USA | 28 | 0 | 81.9 | 25.7 | NA | W | HPLC |
| 33 | 64 | 2001 | RCT | 33 | Vitamin D3 | crystalline form | 25 | 1000 | 150 | Canada | 41.6 | 69.7 | 67.8 | NA | NA | W | RIA |
| 34 | 65 | 2003 | RCT | 15 | Vitamin D3 | Tablets | 25 | 1000 | 160 | USA | 38.7 | 0 | 84.8 | 26.2 | NA | W | HPLC |
| 16 | 66 | 2008 | RCT | 20 | Vitamin D3 | Capsules | 25 | 1000 | 77 | USA | 40 | 65 | NA | 30 | NA | W | LC-MS |
| 11 | 67 | 2013 | RCT | 90 | Vitamin D3 | Capsules | 25 | 1000 | 360 | UK | 64.9 | 100 | 69.4 | 25.2 | NA | N | LC-MS |
| 35 | 68 | 2013 | Non-RCT | 9 | Vitamin D3 | Capsules | 25 | 1000 | 77 | USA | NA | 88.9 | NA | NA | NA | NA | LC-MS |
| 36 | 69 | 2013 | RCT | 81 | Vitamin D3 | Capsules | 25 | 1000 | 84 | USA | 51.1 | 72.8 | NA | 30.5 | NA | N | RIA |
| 19 | 71 | 2015 | RCT | 16 | Vitamin D | NA | 30 | 1200 | 120 | Netherlands | 53 | 0 | NA | 29 | NA | NA | LC-MS |
| 15 | 72 | 2015 | RCT | 14 | Vitamin D3 | Capsules | 30 | 1200 | 112 | China | 34.3 | 50 | NA | 22 | NA | W | LC-MS |
| 37 | 73 | 2011 | RCT | 32 | Vitamin D3 | Capsules | 40 | 1600 | 360 | USA | 77 | 64 | NA | 26.6 | NA | N | HPLC |
| 38 | 75 | 1977 | Non-RCT | 18 | Vitamin D3 | NA | 45 | 1800 | 28 | USA | NA | NA | NA | NA | NA | NA | CPBA |
| 39 | 76 | 1990 | RCT | 25 | Vitamin D3 | Tablets | 45 | 1800 | 77 | Finland | 69.4 | 100 | 70.7 | NA | 0.9 | W | HPLC |
| 39 | 77 | 1990 | RCT | 30 | Vitamin D3 | Tablets | 45 | 1800 | 77 | Finland | 82.2 | 100 | 62.1 | NA | 0.9 | W | HPLC |
| 15 | 78 | 1977 | Non-RCT | 9 | Vitamin D | Tablets | 50 | 2000 | 180 | UK | NA | NA | NA | NA | NA | NA | CPBA |
| 40 | 79 | 1990 | RCT | 30 | Vitamin D3 | Capsules | 50 | 2000 | 42 | USA | 81.1 | 83 | NA | NA | 0.9 | NA | CPBA |
| 41 | 80 | 2009 | RCT | 78 | Vitamin D3 | Tablets | 50 | 2000 | 84 | USA | 59.3 | 78.2 | NA | 26.1 | NA | W | RIA |
| 42 | 81 | 2011 | RCT | 17 | Vitamin D3 | Capsules | 50 | 2000 | 180 | USA | 79.7 | 0 | NA | 26.7 | 1.3 | NA | RIA |
| 43 | 82 | 2012 | RCT | 11 | Vitamin D3 | Tablets | 50 | 2000 | 90 | Australia | 45.5 | 73 | 69.7 | 27 | NA | N | CHEMI |
| 44 | 83 | 2013 | RCT | 42 | Vitamin D3 | Tablets | 50 | 2000 | 56 | Germany | 35.6 | 62 | NA | 24 | 0.9 | W | LC-MS |
| 36 | 84 | 2013 | RCT | 83 | Vitamin D3 | Capsules | 50 | 2000 | 90 | USA | 50.3 | 66.2 | NA | 31.9 | NA | N | RIA |
| 45 | 85 | 2013 | RCT | 50 | Vitamin D3 | NA | 50 | 2000 | 42 | Netherlands | 64 | 4 | NA | NA | NA | N | LC-MS |
| 46 | 87 | 2015 | RCT | 17 | Vitamin D3 | NA | 50 | 2000 | 84 | USA | 36.5 | 100 | NA | 30.4 | NA | N | RIA |
| 13 | 88 | 2015 | RCT | 15 | Vitamin D3 | Capsules | 50 | 2000 | 112 | China | 33.5 | 53 | NA | 22.2 | NA | W | LC-MS |
| 47 | 91 | 2017 | RCT | 16 | Vitamin D3 | Capsules | 60 | 2400 | 112 | USA | 36.9 | NA | NA | 25.7 | NA | N | CHEMI |
| 48 | 94 | 1998 | RCT | 19 | Vitamin D3 | NA | 100 | 4000 | 14 | Canada | 38 | NA | NA | NA | NA | N | RIA |
| 48 | 95 | 1998 | RCT | 18 | Vitamin D3 | NA | 100 | 4000 | 14 | Canada | 38 | NA | NA | NA | NA | N | RIA |
| 48 | 96 | 1998 | RCT | 18 | Vitamin D3 | NA | 100 | 4000 | 14 | Canada | 38 | NA | NA | NA | NA | N | RIA |
| 33 | 97 | 2001 | RCT | 28 | Vitamin D3 | Crystalline form | 100 | 4000 | 150 | Canada | 39.9 | 64.3 | 66.4 | NA | NA | W | RIA |
| 49 | 101 | 2012 | RCT | 35 | Vitamin D3 | NA | 100 | 4000 | 360 | USA | 65.3 | 0 | NA | 28.36 | NA | N | RIA |
| 49 | 102 | 2012 | RCT | 12 | Vitamin D3 | NA | 100 | 4000 | 360 | USA | 63.2 | 0 | NA | 28.4 | NA | N | RIA |
| 50 | 103 | 2013 | RCT | 14 | Vitamin D3 | Capsules | 100 | 4000 | 21 | USA | 31.2 | 56.7 | NA | 24.8 | 0.8 | N | NA |
| 50 | 104 | 2013 | RCT | 15 | Vitamin D3 | Capsules | 100 | 4000 | 21 | USA | 31.9 | 53.3 | NA | 24.6 | 0.8 | N | NA |
| 36 | 105 | 2013 | RCT | 83 | Vitamin D3 | Capsules | 100 | 4000 | 180 | USA | 51.3 | 65.1 | NA | 31.4 | NA | N | RIA |
| 46 | 106 | 2015 | RCT | 18 | Vitamin D3 | NA | 100 | 4000 | 84 | USA | 38.3 | 88.9 | NA | 31.4 | NA | N | RIA |
| 51 | 107 | 2015 | RCT | 14 | Vitamin D | Capsules | 100 | 4000 | 56 | USA | 32.8 | 0 | 78.8 | 23.4 | NA | W | ELISA |
| 51 | 108 | 2015 | RCT | 6 | Vitamin D | Capsules | 100 | 4000 | 56 | USA | 32.2 | 0 | 76.1 | 23.1 | NA | W | ELISA |
| 34 | 112 | 2003 | RCT | 15 | Vitamin D3 | Tablets | 125 | 5000 | 130 | USA | 38.7 | 0 | 84.8 | 26.2 | NA | W | HPLC |
| 52 | 113 | 2009 | Non-RCT | 45 | Vitamin D3 | NA | 125 | 5000 | 360 | Romania | 71 | 62 | NA | NA | NA | N | RIA |
| 53 | 114 | 2011 | RCT | 63 | Vitamin D3 | Capsules | 125 | 5000 | 42 | Australia | 21.45 | 61.9 | NA | NA | NA | NA | LC-MS |
| 54 | 115 | 2012 | RCT | 15 | Vitamin D3 | Tablets | 125 | 5000 | 90 | Australia | 47.4 | 40 | 81 | 28 | NA | N | RIA |
| 55 | 116 | 2013 | Non-RCT | 91 | Vitamin D3 | Capsules | 125 | 5000 | 56 | UK | 23 | 0 | 75 | 24 | NA | W | LC-MS |
| 56 | 117 | 2014 | RCT | 20 | Vitamin D3 | NA | 125 | 5000 | 28 | USA | 25.8 | 100 | 63.4 | 22.8 | NA | N | LC-MS |
| 38 | 119 | 1977 | Non-RCT | 18 | Vitamin D3 | NA | 250 | 10000 | 28 | USA | NA | NA | NA | NA | NA | NA | CPBA |
| 1 | 120 | 1982 | Non-RCT | 8 | Vitamin D3 | arachis oil | 250 | 10000 | 70 | UK | 22.5 | 25 | 45.6 | NA | NA | NA | HPLC |
| 32 | 121 | 1998 | Non-RCT | 10 | Vitamin D3 | Capsules | 250 | 10000 | 56 | USA | 28 | 0 | 81.6 | 25.7 | NA | W | HPLC |
| 34 | 122 | 2003 | RCT | 15 | Vitamin D3 | Tablets | 250 | 10000 | 130 | USA | 38.7 | 0 | 84.8 | 26.2 | NA | W | HPLC |
| 57 | 123 | 2011 | Non-RCT | 8 | Vitamin D3 | Capsules | 250 | 10000 | 28 | USA | 47 | 62.5 | 93 | 30 | NA | NA | LC-MS |
| 38 | 124 | 1977 | Non-RCT | 18 | Vitamin D3 | NA | 250 | 10000 | 28 | USA | NA | NA | NA | NA | NA | NA | CPBA |
| 38 | 125 | 1977 | Non-RCT | 18 | Vitamin D3 | NA | 1000 | 40000 | 28 | USA | NA | NA | NA | NA | NA | NA | CPBA |
| 32 | 126 | 1998 | Non-RCT | 14 | Vitamin D3 | Capsules | 1250 | 50000 | 56 | USA | 28 | 0 | 81.6 | 25.7 | NA | W | HPLC |

* SCR: Serum creatine

§ Season. W: winter after imputation. N: Non-winter after imputation
