## Supplementary material for "Modelling 25-hydroxyvitamin D responses to different recommended daily intakes of vitamin D": Table S2

**Table S2.** References for repeated dose 25(OH)D PK.

| Ref No. | Reference Source |
| --- | --- |
| 1 | Davie, M. W. J., Lawson, D. E. M., Emberson, C., Barnes, J. L. C., Roberts, G. E., & Barnes, N. D. (1982). *Vitamin D from Skin: Contribution to Vitamin D Status Compared with Oral Vitamin D in Normal and Anticonvulsant-Treated Subjects. Clinical Science, 63(5), 461–472.* |
| 2 | LIPS, P., WIERSINGA, A., VAN GINKEL, F. C., JONGEN, M. J. M., NETELENBOS, J. C., HACKENG, W. H. L., … VAN DER VIJGH, W. J. F. (1988). The Effect of Vitamin D Supplementation on Vitamin D Status and Parathyroid Function in Elderly Subjects*. The Journal of Clinical Endocrinology & Metabolism, 67(4), 644–650. |
| 3 | Chel, V. G. M., Ooms, M. E., Popp-Snijders, C., Pavel, S., Schothorst, A. A., Meulemans, C. C. E., & Lips, P. (1998). *Ultraviolet Irradiation Corrects Vitamin D Deficiency and Suppresses Secondary Hyperparathyroidism in the Elderly. Journal of Bone and Mineral Research, 13(8), 1238–1242.* |
| 4 | Grados, F., Brazier, M., Kamel, S., Duver, S., Heurtebize, N., Maamer, M., … Fardellone, P. (2003). *Effects on bone mineral density of calcium and vitamin D supplementation in elderly women with vitamin D deficiency. Joint Bone Spine, 70(3), 203–208.* |
| 5 | Larsen, E. R., Mosekilde, L., & Foldspang, A. (2003). *Vitamin D and Calcium Supplementation Prevents Osteoporotic Fractures in Elderly Community Dwelling Residents: A Pragmatic Population-Based 3-Year Intervention Study. Journal of Bone and Mineral Research, 19(3), 370–378.* |
| 6 | Matsumoto, T., Miki, T., Hagino, H., Sugimoto, T., Okamoto, S., Hirota, T., … Nakamura, T. (2005). *A New Active Vitamin D, ED-71, Increases Bone Mass in Osteoporotic Patients under Vitamin D Supplementation: A Randomized, Double-Blind, Placebo-Controlled Clinical Trial. The Journal of Clinical Endocrinology & Metabolism, 90(9), 5031–5036.* |
| 7 | Pignotti, G. A. P., Genaro, P. S., Pinheiro, M. M., Szejnfeld, V. L., & Martini, L. A. (2009). *Is a lower dose of vitamin D supplementation enough to increase 25(OH)D status in a sunny country? European Journal of Nutrition, 49(5), 277–283.* |
| 8 | Karaplis, A. C., Chouha, F., Djandji, M., Sampalis, J. S., & Hanley, D. A. (2011). *Vitamin D Status and Response to Daily 400 IU Vitamin D3 and Weekly Alendronate 70 mg in Men and Women with Osteoporosis. Annals of Pharmacotherapy, 45(5), 561–568.* |
| 9 | Holvik, K., Madar, A. A., Meyer, H. E., Lofthus, C. M., & Stene, L. C. (2012). *Changes in the vitamin D endocrine system and bone turnover after oral vitamin D3 supplementation in healthy adults: results of a randomised trial. BMC Endocrine Disorders, 12(1).* |
| 10 | Nimitphong, H., Saetung, S., Chanprasertyotin, S., Chailurkit, L., & Ongphiphadhanakul, B. (2013). *Changes in circulating 25-hydroxyvitamin D according to vitamin D binding protein genotypes after vitamin D3 or D2supplementation. Nutrition Journal, 12(1).* |
| 11 | Macdonald, H. M., Wood, A. D., Aucott, L. S., Black, A. J., Fraser, W. D., Mavroeidi, A., … Thies, F. (2013). *Hip bone loss is attenuated with 1000 IU but not 400 IU daily vitamin D3: A 1-year double-blind RCT in postmenopausal women. Journal of Bone and Mineral Research, 28(10), 2202–2213.* |
| 12 | Wagner, D., Trudel, D., Van der Kwast, T., Nonn, L., Giangreco, A. A., Li, D., … Vieth, R. (2013). *Randomized Clinical Trial of Vitamin D3Doses on Prostatic Vitamin D Metabolite Levels and Ki67 Labeling in Prostate Cancer Patients. The Journal of Clinical Endocrinology & Metabolism, 98(4), 1498–1507.* |
| 13 | Yao, P., Lu, L., Hu, Y., Liu, G., Chen, X., Sun, L., … Lin, X. (2015). *A dose–response study of vitamin D3 supplementation in healthy Chinese: a 5-arm randomized, placebo-controlled trial. European Journal of Nutrition, 55(1), 383–392.* |
| 14 | Somerville, P. J., Lien, J. W. K., & Kaye, M. (1977). The Calcium and Vitamin D Status in An Elderly Female Population and Their Response to Administered Supplemental Vitamin D3. Journal of Gerontology, 32(6), 659–663. |
| 15 | MacLennan, W. J., & Hamilton, J. C. (1977). Vitamin D supplements and 25-hydroxy vitamin D concentrations in the elderly. BMJ, 2(6091), 859–861. doi:10.1136/bmj.2.6091.860 |
| 16 | Holick, M. F., Biancuzzo, R. M., Chen, T. C., Klein, E. K., Young, A., Bibuld, D., … Tannenbaum, A. D. (2008). *Vitamin D2Is as Effective as Vitamin D3in Maintaining Circulating Concentrations of 25-Hydroxyvitamin D. The Journal of Clinical Endocrinology & Metabolism, 93(3), 677–681.* |
| 17 | Hiremath, V. P., Rao, C. B., Naik, V., & Prasad, K. V. (2013). Anti‐inflammatory effect of vitamin D on gingivitis: A dose‐response randomised control trial. Oral Health & Preventive Dentistry, 11, 61–69. https://doi.org/10.3290/j.ohpd.a29377 |
| 18 | Chel, V., Wijnhoven, H. A. H., Smit, J. H., Ooms, M., & Lips, P. (2007). Efficacy of different doses and time intervals of oral vitamin D supplementation with or without calcium in elderly nursing home residents. Osteoporosis International, 19(5), 663–671. |
| 19 | Heijboer, A. C., Oosterwerff, M., Schroten, N. F., Eekhoff, E. M. W., Chel, V. G. M., de Boer, R. A., … Lips, P. (2015). Vitamin D supplementation and testosterone concentrations in male human subjects. Clinical Endocrinology, 83(1), 105–110. |
| 20 | Van Der Klis, F. R. M., Jonxis, J. H. P., Van Doormaal, J. J., Sikkens, P., Saleh, A. E. C., & Muskiet, F. A. J. (1996). Changes in vitamin-D metabolites and parathyroid hormone in plasma following cholecalciferol administration to pre- and postmenopausal women in the Netherlands in early spring and to postmenopausal women in Curaçao. British Journal of Nutrition, 75(04), 637. |
| 21 | Kyriakidou-Himonas, M., Aloia, J. F., & Yeh, J. K. (1999). Vitamin D Supplementation in Postmenopausal Black Women. The Journal of Clinical Endocrinology & Metabolism, 84(11), 3988–3990. |
| 22 | Hunter D, Major P, Arden N, Swaminathan R, Andrew T, Mac Gregor AJ, Keen R, Snieder H & Spector TD (2000) A ran domized controlled trial of vitamin D supplementation onpreventing postmenopausal bone loss and modifying bone metabolism using identical twin pairs. Journal of Bone and Mineral Research 15, 276–283. |
| 23 | Patel, R., Collins, D., Bullock, S., Swaminathan, R., Blake, G. M., & Fogelman, I. (2001). The Effect of Season and Vitamin D Supplementation on Bone Mineral Density in Healthy Women: A Double-Masked Crossover Study. Osteoporosis International, 12(4), 319–325. |
| 24 | Harris, S. S., & Dawson-Hughes, B. (2002). Plasma Vitamin D and 25OHD Responses of Young and Old Men to Supplementation with Vitamin D3. Journal of the American College of Nutrition, 21(4), 357–362. |
| 25 | Bischoff, H. A., Stähelin, H. B., Dick, W., Akos, R., Knecht, M., Salis, C., … Conzelmann, M. (2003). Effects of Vitamin D and Calcium Supplementation on Falls: A Randomized Controlled Trial. Journal of Bone and Mineral Research, 18(2), 343–351. |
| 26 | Talwar SA, Aloia JF, Pollack S, Yeh JK. Dose response to vitamin D supplementation among postmenopausal African American women. Am J Clin Nutr. 2007;86:1657–1662. |
| 27 | KUWABARA, A., TSUGAWA, N., TANAKA, K., FUJII, M., KAWAI, N., MUKAE, S., … OKANO, T. (2009). Improvement of Vitamin D Status in Japanese Institutionalized Elderly by Supplementation with 800 IU of Vitamin D3. Journal of Nutritional Science and Vitaminology, 55(6), 453–458. doi:10.3177/jnsv.55.453 |
| 28 | Wicherts, I. S., Boeke, A. J. P., van der Meer, I. M., van Schoor, N. M., Knol, D. L., & Lips, P. (2010). Sunlight exposure or vitamin D supplementation for vitamin D-deficient non-western immigrants: a randomized clinical trial. Osteoporosis International, 22(3), 873–882. |
| 29 | Bischoff-Ferrari, H. A., Dawson-Hughes, B., Stöcklin, E., Sidelnikov, E., Willett, W. C., Edel, J. O., … Egli, A. (2011). Oral supplementation with 25(OH)D3versus vitamin D3: Effects on 25(OH)D levels, lower extremity function, blood pressure, and markers of innate immunity. Journal of Bone and Mineral Research, 27(1), 160–169. |
| 30 | Cashman, K. D., Seamans, K. M., Lucey, A. J., Stöcklin, E., Weber, P., Kiely, M., & Hill, T. R. (2012). Relative effectiveness of oral 25-hydroxyvitamin D3 and vitamin D3 in raising wintertime serum 25-hydroxyvitamin D in older adults. The American Journal of Clinical Nutrition, 95(6), 1350–1356. |
| 31 | Wijnen, H., Salemink, D., Roovers, L., Taekema, D., & de Boer, H. (2015). Vitamin D Supplementation in Nursing Home Patients: Randomized Controlled Trial of Standard Daily Dose Versus Individualized Loading Dose Regimen. Drugs & Aging, 32(5), 371–378. |
| 32 | Barger-Lux, M. J., Heaney, R. P., Dowell, S., Chen, T. C., & Holick, M. F. (1998). Vitamin D and its Major Metabolites: Serum Levels after Graded Oral Dosing in Healthy Men. Osteoporosis International, 8(3), 222–230. |
| 33 | Vieth R, Chan PC, MacFarlane GD. Efficacy and safety of vitamin D3 intake exceeding the lowest observed adverse effect level. Am J Clin Nutr 2001;73:288 –94. |
| 34 | Heaney, R. P., Davies, K. M., Chen, T. C., Holick, M. F., & Barger-Lux, M. J. (2003). Human serum 25-hydroxycholecalciferol response to extended oral dosing with cholecalciferol. The American Journal of Clinical Nutrition, 77(1), 204–210. |
| 35 | Biancuzzo, R. M., Clarke, N., Reitz, R. E., Travison, T. G., & Holick, M. F. (2013). Serum Concentrations of 1,25-Dihydroxyvitamin D2and 1,25-Dihydroxyvitamin D3in Response to Vitamin D2and Vitamin D3Supplementation. The Journal of Clinical Endocrinology & Metabolism, 98(3), 973–979. |
| 36 | Ng, K., Scott, J. B., Drake, B. F., Chan, A. T., Hollis, B. W., Chandler, P. D., … Fuchs, C. S. (2013). Dose response to vitamin D supplementation in African Americans: results of a 4-arm, randomized, placebo-controlled trial. The American Journal of Clinical Nutrition, 99(3), 587–598. |
| 37 | Binkley, N., Gemar, D., Engelke, J., Gangnon, R., Ramamurthy, R., Krueger, D., & Drezner, M. K. (2011). *Evaluation of Ergocalciferol or Cholecalciferol Dosing, 1,600 IU Daily or 50,000 IU Monthly in Older Adults. The Journal of Clinical Endocrinology & Metabolism, 96(4), 981–988.* |
| 38 | Stamp, T. C. B., Haddad, J. G., & Twigg, C. A. (1977). *COMPARISON OF ORAL 25-HYDROXYCHOLECALCIFEROL, VITAMIN D, AND ULTRAVIOLET LIGHT AS DETERMINANTS OF CIRCULATING 25-HYDROXYVITAMIN D. The Lancet, 309(8026), 1341–1343.* |
| 39 | Honkanen, R., Alhava, E., Parviainen, M., Talasniemi, S., & Mönkkönen, R. (1990). The Necessity and Safety of Calcium and Vitamin D in the Elderly. Journal of the American Geriatrics Society, 38(8), 862–866. doi:10.1111/j.1532-5415. |
| 40 | Himmelstein, S., Clemens, T. L., Rubin, A., & Lindsay, R. (1990). Vitamin D supplementation in elderly nursing home residents increases 25(OH)D but not 1,25(OH)2D. The American Journal of Clinical Nutrition, 52(4), 701–706. |
| 41 | LI-NG, M., ALOIA, J. F., POLLACK, S., CUNHA, B. A., MIKHAIL, M., YEH, J., & BERBARI, N. (2009). *A randomized controlled trial of vitamin D3 supplementation for the prevention of symptomatic upper respiratory tract infections. Epidemiology and Infection, 137(10), 1396.* |
| 42 | Cherniack, E. P., Florez, H. J., Hollis, B. W., Roos, B. A., Troen, B. R., & Levis, S. (2011). *The Response of Elderly Veterans to Daily Vitamin D3 Supplementation of 2,000 IU: A Pilot Efficacy Study. Journal of the American Geriatrics Society, 59(2), 286–290.* |
| 43 | Diamond, T., Wong, Y. K., & Golombick, T. (2012). *Effect of oral cholecalciferol 2,000 versus 5,000 IU on serum vitamin D, PTH, bone and muscle strength in patients with vitamin D deficiency. Osteoporosis International, 24(3), 1101–1105.* |
| 44 | Lehmann, U., Hirche, F., Stangl, G. I., Hinz, K., Westphal, S., & Dierkes, J. (2013). *Bioavailability of Vitamin D2and D3in Healthy Volunteers, a Randomized Placebo-Controlled Trial. The Journal of Clinical Endocrinology & Metabolism, 98(11), 4339–4345.* |
| 45 | Schroten, N. F., Ruifrok, W. P. T., Kleijn, L., Dokter, M. M., Silljé, H. H., Lambers Heerspink, H. J., … de Boer, R. A. (2013). Short-term vitamin D3 supplementation lowers plasma renin activity in patients with stable chronic heart failure: An open-label, blinded end point, randomized prospective trial (VitD-CHF trial). American Heart Journal, 166(2), 357–364.e2. |
| 46 | Sorva, A., Risteli, J., Risteli, L., Välimäki, M., & Tilvis, R. (1991). Effects of vitamin D and calcium on markers of bone metabolism in geriatric patients with low serum 25-hydroxyvitamin D levels. Calcified Tissue International, 49(S1), S88–S89. |
| 47 | Shieh, A., Ma, C., Chun, R. F., Witzel, S., Rafison, B., Contreras, H. T. M., … Adams, J. S. (2017). Effects of Cholecalciferol vs Calcifediol on Total and Free 25-Hydroxyvitamin D and Parathyroid Hormone. The Journal of Clinical Endocrinology & Metabolism, 102(4), 1133–1140. |
| 48 | Trang, H. M., Cole, D. E., Rubin, L. A., Pierratos, A., Siu, S., & Vieth, R. (1998). *Evidence that vitamin D3 increases serum 25-hydroxyvitamin D more efficiently than does vitamin D2. The American Journal of Clinical Nutrition, 68(4), 854–858.* |
| 49 | Garrett-Mayer, E., Wagner, C. L., Hollis, B. W., Kindy, M. S., & Gattoni-Celli, S. (2012). *Vitamin D3 supplementation (4000 IU/d for 1 y) eliminates differences in circulating 25-hydroxyvitamin D between African American and white men. The American Journal of Clinical Nutrition, 96(2), 332–336.* |
| 50 | Hata, T. R., Audish, D., Kotol, P., Coda, A., Kabigting, F., Miller, J., … Gallo, R. L. (2013). *A randomized controlled double-blind investigation of the effects of vitamin D dietary supplementation in subjects with atopic dermatitis. Journal of the European Academy of Dermatology and Venereology, 28(6), 781–789.* |
| 51 | Scholten, S., Sergeev, I., Birger, C., & Song, Q. (2015). *Effects of vitamin D and quercetin, alone and in combination, on cardiorespiratory fitness and muscle function in physically active male adults. Open Access Journal of Sports Medicine, 229.* |
| 52 | Mocanu, V., Stitt, P. A., Costan, A. R., Voroniuc, O., Zbranca, E., Luca, V., & Vieth, R. (2009). *Long-term effects of giving nursing home residents bread fortified with 125 μg (5000 IU) vitamin D3 per daily serving. The American Journal of Clinical Nutrition, 89(4), 1132–1137.* |
| 53 | Dean, A. J., Bellgrove, M. A., Hall, T., Phan, W. M. J., Eyles, D. W., Kvaskoff, D., & McGrath, J. J. (2011). *Effects of Vitamin D Supplementation on Cognitive and Emotional Functioning in Young Adults – A Randomised Controlled Trial. PLoS ONE, 6(11), e25966.* |
| 54 | Diamond, T., Wong, Y. K., & Golombick, T. (2012). *Effect of oral cholecalciferol 2,000 versus 5,000 IU on serum vitamin D, PTH, bone and muscle strength in patients with vitamin D deficiency. Osteoporosis International, 24(3), 1101–1106.* |
| 55 | Close, G. L., Russell, J., Cobley, J. N., Owens, D. J., Wilson, G., Gregson, W., … Morton, J. P. (2013). Assessment of vitamin D concentration in non-supplemented professional athletes and healthy adults during the winter months in the UK: implications for skeletal muscle function. Journal of Sports Sciences, 31(4), 344–353. |
| 56 | Meekins, M. E., Oberhelman, S. S., Lee, B. R., Gardner, B. M., Cha, S. S., Singh, R. J., … Thacher, T. D. (2014). *Pharmacokinetics of daily versus monthly vitamin D3 supplementation in non-lactating women. European Journal of Clinical Nutrition, 68(5), 632–634.* |
| 57 | Nazarian, S., St. Peter, J. V., Boston, R. C., Jones, S. A., & Mariash, C. N. (2011). *Vitamin D3 supplementation improves insulin sensitivity in subjects with impaired fasting glucose. Translational Research, 158(5), 276–281.* |
