## Supplementary material for "Modelling 25-hydroxyvitamin D responses to different recommended daily intakes of vitamin D": Table S3 and S4

**Table S3.** Model development for vitamin D pharmacokinetics.

| **Model no** | **No of trial arms** | **Fixed Effects** | **Random Effects** $\eta$ | **Covariate** | **AIC** | **BIC** | **OBJFV** | **Comments** | **Fitting results** |
| --- | --- | --- | --- | --- | --- | --- | --- | --- | --- |
| **1** | 13 | CL_MAX_ | CL_MAX_ | - | 480.5137 | 490.2370 | 318.1321 | Good inference of CL_MAX_ | Fit.s1 |
| **2** | 13 | Kp_rb_ | Kp_rb_ | - | 576.0267 | 585.7500 | 413.6451 | Goodness of fit is worse than model 1  Shrinkage of Kp_rb_ = 94.1%, indicating overfitting | Fit.s2 |
| **3** | 13 | k_a_ | k_a_ | - | 523.3436 | 533.0668 | 360.9619 | Goodness of fit is worse than model 1  Shrinkage of k_a_ = 40.0%, indicating overfitting | Fit.s3 |
| **4** | 13 | CL_MAX_, Kp_rb_ | CL_MAX_ and Kp_rb_ are  assumed independent | - | 469.3641 | 483.9490 | 302.9824 | Shrinkage of Kp_rb_ = 96.2%, indicating overfitting | Fit.s4 |
| **5** | 13 | CL_MAX_, k_a_ | CL_MAX_ and k_a_ are  assumed independent | - | 464.5561 | 479.1410 | 298.1744 | Shrinkage of k_a_ = 91.4%, indicating overfitting | Fit.s5 |
| **6** | 13 | CL_MAX_, Kp_rb_, k_a_ | CL_MAX_,  Kp_rb_ and k_a_ are  assumed independent | - | 6564.5832 | 6584.0297 | 6394.2015 | Goodness of fit is much worse than model 1, indicating overfitting | Fit.s6 |
| **7** | 13 | CL_MAX_,  Kp_rb_ and Kp_l_ | CL_MAX_,  Kp_rb_ and Kp_l_ are assumed independent | - | 460.6877 | 480.1342 | 290.3060 | Kp_rb_ and Kp_l_ have shrinkage over 80%, indicating overfitting | Fit.s7 |
| **8** | 13 | CL_MAX_, k_a_, Kp_rb_ and Kp_l_ | CL_MAX_, k_a_, Kp_rb_ and Kp_l_ are assumed independent | - | 906.5595 | 930.8677 | 732.1779 | Goodness of fit is much worse than model 1, indicating overfitting | Fit.s8 |
| **9** | 13 | CL_MAX_ and  Kp_rb_ | IIV: CL_MAX_ and Kp_rb_ are assumed to independent  IOV: CL_MAX_ for single dose and repeated doses | - | 476.3317 | 493.3474 | 307.9501 | Goodness of fit is worse than model 1 | Fit.s9 |
| **10** | 13 | CL_MAX_, k_a_, and Kp_rb_ | CL_MAX_ and Kp_rb_ are  Assumed correlated, while k_a_ is assumed independent from the other two | - | 2132.0954 | 2153.9727 | 1959.7137 | Goodness of fit is much worse than model 7, indicating overfitting | Fit.s10 |
| **11** | 13 | CL_MAX_ | IIV: CL_MAX_  IOV: CL_MAX_ for single dose and repeated doses | - | 472.0470 | 484.2011 | 307.6653 | Goodness of fit is better than model 1  Good inference is made for both IIV and IOV  **This is the best model** | Fit.s11 |

**Table S4.** Model development for 25(OH)D pharmacokinetics.

| **Model no** | **No of trial arms** | **Fixed Effects** | **Random Effects** | **Covariate** | **AIC** | **BIC** | **OBJFV** | **Comments** | **Fitting results** |
| --- | --- | --- | --- | --- | --- | --- | --- | --- | --- |
| **1** | 90 | CL_MAX_ | CL_MAX_ | - | 2181.3625 | 2196.0968 | 1633.0266 |  | Fit.s1.25OHD |
| **2** | 90 | Kp25_rb_ | Kp25_rb_ | - | 2025.9436 | 2040.6779 | 1477.6078 | Goodness of fit is improved compared with model 1 | Fit.s2.25OHD |
| **3** | 33 | Kp25_rb_ | Kp25_rb_ | - | 756.2609 | 766.7608 | 560.7975 | Fitted to trial arms with bodyweight reported | Fit.s3.25OHD |
| **4** | 33 | Kp25_rb_ | Kp25_rb_ | Weight for Kp25_rb_ | 748.5902 | 761.7150 | 551.1267 | Introducing bodyweight as a covariate only marginally improved the goodness of fit | Fit.s4.25OHD |
| **5** | 90 | C_50_ | C_50_ | - | 2260.3839 | 2275.1182 | 1712.0480 | Goodness of fit is worse than model 2 | Fit.s5.25OHD |
| **6** | 90 | CL_MAX_ and  Kp25_rb_ | CL_MAX_ and  Kp25_rb_ are  assumed independent | - | 1898.1484 | 1920.2499 | 1345.8126 | Goodness of fit is better than model 2 | Fit.s6.25OHD |
| **7** | 90 | CL_MAX_ and  Kp25_rb_ | CL_MAX_ and Kp25_rb_ are assumed to be correlated | - | 1874.7876 | 1900.5727 | 1320.4518 | Goodness of fit is better than model 6  **This is the best model without any covariate (e.g. if bodyweight is unknown).** | Fit.s7.25OHD |
| **8** | 33 | CL_MAX_ and  Kp25_rb_ | CL_MAX_ and Kp25_rb_ are assumed to be correlated | - | 701.6326 | 720.0074 | 500.1691 | Fitted to trial arms with bodyweight reported | Fit.s8.25OHD |
| **9** | 33 | CL_MAX_ and  Kp25_rb_ | CL_MAX_ and Kp25_rb_ are assumed to be correlated | Weight for Kp25_rb_ only | 695.0449 | 716.0447 | 491.5815 | Introducing bodyweight as a covariate marginally improved the goodness of fit | Fit.s9.25OHD |
| **10** | 33 | CL_MAX_ and Kp25_rb_ | CL_MAX_ and Kp25_rb_ are assumed to be correlated | - | 668.9746 | 687.3494 | 467.5111 | Assume volume of distribution is proportional to bodyweight. This improved goodness of fit  **This is the best model if bodyweight is known.** | Fit.s10.25OHD |

^*^Four compartments are considered: venous blood, arterial blood, liver, rest of body.
