## Supplementary material for "Modelling 25-hydroxyvitamin D responses to different recommended daily intakes of vitamin D": Table S5

### Vitamin D Population PBPK Modelling

#### Table 1 Running models

| Run No. | AIC | BIC | OFV |
|---|---|---|---|
| Model 1 | 480.5137 | 490.2370 | 318.1321 |
| Model 2 | 576.0267 | 585.7500 | 413.6451 |
| Model 3 | 523.3436 | 533.0668 | 360.9619 |
| Model 4 | 469.3641 | 483.9490 | 302.9824 |
| Model 5 | 464.5561 | 479.1410 | 298.1744 |
| Model 6 | 6564.5832 | 6584.0297 | 6394.2015 |
| Model 7 | 460.6877 | 480.1342 | 290.3060 |
| Model 8 | 906.5595 | 930.8677 | 732.1779 |
| Model 9 | 476.3317 | 493.3474 | 307.9501 |
| Model 10 | 2132.0954 | 2153.9727 | 1959.7137 |
| Model 11 | 472.0470 | 484.2011 | 307.6653 |

#### Table list. Parameter estimation

Table 1: fit.s1.rds

| | Parameter | Est. | SE | %RSE | Back-transformed(95%CI) | BSV(CV%) | Shrink(SD)% |
|---|---|---|---|---|---|---|---|
| lcl | Clearance (L/h) | -1.27 | 0.14 | 11 | 0.28 (0.213, 0.369) | 46.9 | 7.30%< |
| add.err | | 0.000374 | | | 0.000374 | | |
| prop.err | | 0.195 | | | 0.195 | | |

Table 2: fit.s2.rds

| | Parameter | Est. | SE | %RSE | Back-transformed(95%CI) | BSV(CV%) | Shrink(SD)% |
|---|---|---|---|---|---|---|---|
| lkprb | Partition coefficient for the rest of the body | -2.11 | 0.195 | 9.25 | 0.122 (0.0832, 0.178) | 3.73 | 94.1%> |
| prop.err | | 0.306 | | | 0.306 | | |
| add.err | | 0.000438 | | | 0.000438 | | |

Table 3: fit.s3.rds

| | Parameter | Est. | SE | %RSE | Back-transformed(95%CI) | BSV(CV%) | Shrink(SD)% |
|---|---|---|---|---|---|---|---|
| lka | Absorption rate (/h) | -2.31 | 0.261 | 11.3 | 0.0993 (0.0595, 0.166) | 48.6 | 40.0%> |
| prop.err | | 0.283 | | | 0.283 | | |
| add.err | | 0.000427 | | | 0.000427 | | |

Table 4: fit.s4.rds

| | Parameter | Est. | SE | %RSE | Back-transformed(95%CI) | BSV(CV%) | Shrink(SD)% |
|---|---|---|---|---|---|---|---|
| lkprb | Partition coefficient for the rest of the body | -2.44 | 0.155 | 6.35 | 0.0871 (0.0643, 0.118) | 1.55 | 96.2%> |
| lcl | Clearance (L/h) | -1.27 | 0.139 | 11 | 0.281 (0.214, 0.369) | 46.5 | 7.41%< |
| add.err | | 0.000405 | | | 0.000405 | | |
| prop.err | | 0.194 | | | 0.194 | | |

Table 5: fit.s5.rds

| | Parameter | Est. | SE | %RSE | Back-transformed(95%CI) | BSV(CV%) | Shrink(SD)% |
|---|---|---|---|---|---|---|---|
| lka | Absorption rate (/h) | -1.87 | 0.116 | 6.2 | 0.154 (0.123, 0.193) | 2.61 | 91.4%> |
| lcl | Clearance (L/h) | -1.26 | 0.138 | 11 | 0.283 (0.216, 0.371) | 45.7 | 7.81%< |
| add.err | | 0.000436 | | | 0.000436 | | |
| prop.err | | 0.196 | | | 0.196 | | |

Table 6: fit.s6.rds

| | Parameter | Est. | SE | %RSE | Back-transformed(95%CI) | BSV(CV%) | Shrink(SD)% |
|---|---|---|---|---|---|---|---|
| lka | Absorption rate (/h) | -2.55 | 0.254 | 9.95 | 0.0778 (0.0473, 0.128) | 0.388 | 95.2%> |
| lkprb | Partition coefficient for the rest of the body | -6.41 | 16.3 | 254 | 0.00165 (2.26e-17, 1.2e+11) | 53.5 | 95.6%> |
| lcl | Clearance (L/h) | -1.25 | 0.137 | 10.9 | 0.286 (0.219, 0.374) | 46.6 | 5.40%< |
| add.err | | 0.000399 | | | 0.000399 | | |
| prop.err | | 0.181 | | | 0.181 | | |

‘

Table 7: fit.s7.rds

| | Parameter | Est. | SE | %RSE | Back-transformed(95%CI) | BSV(CV%) | Shrink(SD)% |
|---|---|---|---|---|---|---|---|
| lkpl | Partition coefficient for liver | -0.412 | 20 | 4.84e+03 | 0.662 (6.88e-18, 6.38e+16) | 9.86 | 94.7%> |
| lkprb | Partition coefficient for the rest of the body | -2.37 | 4.12 | 174 | 0.0937 (2.9e-05, 303) | 10.1 | 85.6%> |
| lcl | Clearance (L/h) | -1.27 | 0.138 | 10.9 | 0.28 (0.213, 0.367) | 46.1 | 6.74%< |
| add.err | | 0.000434 | | | 0.000434 | | |
| prop.err | | 0.193 | | | 0.193 | | |

Table 8: fit.s8.rds

| | Parameter | Est. | SE | %RSE | Back-transformed(95%CI) | BSV(CV%) | Shrink(SD)% |
|---|---|---|---|---|---|---|---|
| lka | Absorption rate (/h) | -2.51 | 0.509 | 20.2 | 0.0809 (0.0298, 0.219) | 2.25 | 88.9%> |
| lkpl | Partition coefficient for liver | -6.73 | 607 | 9.01e+03 | 0.00119 (0, inf) | 86.3 | 94.3%> |
| lkprb | Partition coefficient for the rest of the body | -3.65 | 2.11 | 57.8 | 0.0259 (0.000413, 1.62) | 6.05 | 93.8%> |
| lcl | Clearance (L/h) | -1.25 | 0.143 | 11.5 | 0.286 (0.216, 0.379) | 48.6 | 5.41%< |
| add.err | | 0.000421 | | | 0.000421 | | |
| prop.err | | 0.179 | | | 0.179 | | |

Table 9: fit.s9.rds

| | Parameter | Est. | SE | %RSE | Back-transformed(95%CI) | BSV(CV%) | Shrink(SD)% |
|---|---|---|---|---|---|---|---|
| lkprb | Partition coefficient for the rest of the body | -2.59 | 0.171 | 6.6 | 0.0754 (0.0539, 0.105) | 2.90 | 96.2%> |
| lcl | Clearance (L/h) | -1.53 | 0.108 | 7.09 | 0.217 (0.176, 0.268) | 24.5 | 17.7%< |
| lcl_iov1 | | 0.782 | 0.182 | 23.2 | 0.782 (0.426, 1.14) | | |
| add.err | | 0.000447 | | | 0.000447 | | |
| prop.err | | 0.193 | | | 0.193 | | |

Table 10: fit.s10.rds

| | Parameter | Est. | SE | %RSE | Back-transformed(95%CI) | BSV(CV%) | Shrink(SD)% |
|---|---|---|---|---|---|---|---|
| lka | Absorption rate (/h) | -2.53 | 0.391 | 15.5 | 0.0796 (0.037, 0.171) | 0.744 | 95.8%> |
| lkprb | Partition coefficient for the rest of the body | -5.66 | 7.8 | 138 | 0.0035 (7.96e-10, 1.54e+04) | 52.1 | 6.14%< |
| lcl | Clearance (L/h) | -1.26 | 0.138 | 11 | 0.284 (0.217, 0.372) | 47.2 | 5.19%< |
| add.err | | 0.000388 | | | 0.000388 | | |
| prop.err | | 0.18 | | | 0.18 | | |

Table 11: fit.s11.rds

| | Parameter | Est. | SE | %RSE | Back-transformed(95%CI) | BSV(CV%) | Shrink(SD)% |
|---|---|---|---|---|---|---|---|
| lcl | Clearance (L/h) | -1.51 | 0.108 | 7.17 | 0.222 (0.179, 0.274) | 24.3 | 18.0%< |
| lcl_iov1 | IOV constant | 0.711 | 0.183 | 25.7 | 0.711 (0.353, 1.07) | | |
| add.err | | 0.000419 | | | 0.000419 | | |
| prop.err | | 0.195 | | | 0.195 | | |
