## Supplementary material for "Modelling 25-hydroxyvitamin D responses to different recommended daily intakes of vitamin D": Table S6

### 25(OH)D Population PBPK Modelling

#### Table 1 Running models

| i | Model.structure | AIC | BIC | OBJFV |
|---|---|---|---|---|
| 1 | Model 1 | 2181.3625 | 2196.0968 | 1633.0266 |
| 2 | Model 2 | 2025.9436 | 2040.6779 | 1477.6078 |
| 3 | Model 3 | 756.2609 | 766.7608 | 560.7975 |
| 4 | Model 4 | 748.5902 | 761.7150 | 551.1267 |
| 5 | Model 5 | 2260.3839 | 2275.1182 | 1712.0480 |
| 6 | Model 6 | 1898.1484 | 1920.2499 | 1345.8126 |
| 7 | Model 7 | 1874.7876 | 1900.5727 | 1320.4518 |
| 8 | Model 8 | 701.6326 | 720.0074 | 500.1691 |
| 9 | Model 9 | 695.0449 | 716.0447 | 491.5815 |
| 10 | Model 10 | 668.9746 | 687.3494 | 467.5111 |

#### Table list. Parameter estimation

Table 1: fit.s1.25OHD.rds

| | Est. | SE | %RSE | Back-transformed(95%CI) | BSV(CV%) | Shrink(SD)% |
|---|---|---|---|---|---|---|
| TCL_EMAX | -3.03 | 0.0999 | 3.3 | 0.0483 (0.0397, 0.0587) | 48.0 | 50.6%> |
| add.err | 8.89 | | | 8.89 | | |
| prop.err | 0.0299 | | | 0.0299 | | |

Table 2: fit.s2.25OHD.rds

| | Est. | SE | %RSE | Back-transformed(95%CI) | BSV(CV%) | Shrink(SD)% |
|---|---|---|---|---|---|---|
| TKp25rb | -1.18 | 0.143 | 12.2 | 0.308 (0.233, 0.408) | 147. | 18.3%< |
| add.err | 0.00299 | | | 0.00299 | | |
| prop.err | 0.0957 | | | 0.0957 | | |

Table 3: fit.s3.25OHD.rds

| | Est. | SE | %RSE | Back-transformed(95%CI) | BSV(CV%) | Shrink(SD)% |
|---|---|---|---|---|---|---|
| TKp25rb | -1.1 | 0.24 | 21.8 | 0.334 (0.209, 0.534) | 150. | 19.8%< |
| add.err | 0.00317 | | | 0.00317 | | |
| prop.err | 0.11 | | | 0.11 | | |

Table 4: fit.s4.25OHD.rds

| | Est. | SE | %RSE | Back-transformed(95%CI) | BSV(CV%) | Shrink(SD)% |
|---|---|---|---|---|---|---|
| TKp25rb | -1.03 | 0.195 | 18.9 | 0.357 (0.244, 0.523) | 105. | 22.3%= |
| wt.eff | 3.04 | 0.968 | 31.9 | 3.04 (1.14, 4.94) | | |
| add.err | 0.00317 | | | 0.00317 | | |
| prop.err | 0.109 | | | 0.109 | | |

Table 5: fit.s5.25OHD.rds

| | Est. | SE | %RSE | Back-transformed(95%CI) | BSV(CV%) | Shrink(SD)% |
|---|---|---|---|---|---|---|
| TCL_C50 | 4.35 | 0.0729 | 1.68 | 77.3 (67, 89.2) | 9.72 | 86.3%> |
| add.err | 7.98 | | | 7.98 | | |
| prop.err | 0.119 | | | 0.119 | | |

Table 6: fit.s6.25OHD.rds

| | Est. | SE | %RSE | Back-transformed(95%CI) | BSV(CV%) | Shrink(SD)% |
|---|---|---|---|---|---|---|
| TKp25rb | -1.95 | 0.225 | 11.5 | 0.142 (0.0914, 0.221) | 219. | 29.0%= |
| TCL_EMAX | -2.95 | 0.0889 | 3.01 | 0.0522 (0.0439, 0.0622) | 82.7 | 18.4%< |
| add.err | 0.00134 | | | 0.00134 | | |
| prop.err | 0.0562 | | | 0.0562 | | |

Table 7: fit.s7.25OHD.rds

| | Est. | SE | %RSE | Back-transformed(95%CI) | BSV(CV%) | Shrink(SD)% |
|---|---|---|---|---|---|---|
| TKp25rb | -2.08 | 0.232 | 11.1 | 0.124 (0.0789, 0.196) | 244. | 24.6%= |
| TCL_EMAX | -2.9 | 0.0848 | 2.92 | 0.055 (0.0466, 0.065) | 79.0 | 13.4%< |
| add.err | 0.00105 | | | 0.00105 | | |
| prop.err | 0.0551 | | | 0.0551 | | |

‘

Table 8: fit.s8.25OHD.rds

| | Est. | SE | %RSE | Back-transformed(95%CI) | BSV(CV%) | Shrink(SD)% |
|---|---|---|---|---|---|---|
| TKp25rb | -1.79 | 0.399 | 22.2 | 0.167 (0.0762, 0.364) | 237. | 30.1%> |
| TCL_EMAX | -3 | 0.104 | 3.47 | 0.0499 (0.0407, 0.0611) | 52.3 | 26.0%= |
| add.err | 0.00165 | | | 0.00165 | | |
| prop.err | 0.0623 | | | 0.0623 | | |

Table 9: fit.s9.25OHD.rds

| | Est. | SE | %RSE | Back-transformed(95%CI) | BSV(CV%) | Shrink(SD)% |
|---|---|---|---|---|---|---|
| TKp25rb | -1.58 | 0.313 | 19.9 | 0.206 (0.112, 0.381) | 156. | 27.6%= |
| TCL_EMAX | -3.01 | 0.111 | 3.67 | 0.0492 (0.0396, 0.0611) | 52.5 | 25.7%= |
| wt.eff | 2.62 | 1.32 | 50.4 | 2.62 (0.0333, 5.21) | | |
| add.err | 0.00296 | | | 0.00296 | | |
| prop.err | 0.0629 | | | 0.0629 | | |

Table 10: fit.s10.25OHD.rds

| | Est. | SE | %RSE | Back-transformed(95%CI) | BSV(CV%) | Shrink(SD)% |
|---|---|---|---|---|---|---|
| TKp25rb | -1.35 | 0.249 | 18.4 | 0.258 (0.158, 0.421) | 120. | 27.6%= |
| TCL_EMAX | -3.04 | 0.113 | 3.7 | 0.0477 (0.0383, 0.0595) | 58.1 | 20.2%= |
| add.err | 0.000794 | | | 0.000794 | | |
| prop.err | 0.046 | | | 0.046 | | |
